# A linear Mixed Model to Estimate COVID-19-induced Excess Mortality

**DOI:** 10.1101/2021.05.10.21256942

**Authors:** Johan Verbeeck, Christel Faes, Thomas Neyens, Niel Hens, Geert Verbeke, Patrick Deboosere, Geert Molenberghs

**Affiliations:** Data Science Institute (DSI), Interuniversity Institute for Biostatistics and statistical Bioinformatics (I-BioStat), Hasselt University, BE-3500 Hasselt, Belgium; Interuniversity Institute for Biostatistics and statistical Bioinformatics (I-BioStat), KULeuven, BE-3000 Leuven, Belgium; Centre for Health Economics Research and Modelling of Infectious Diseases (CHERMID), Vaccine & Infectious Disease Institute (VAXINFECTIO), University of Antwerp, BE-2000 Antwerp, Belgium; Interface Demography (ID), Department of Sociology, Vrije Universiteit Brussel, BE-1050 Brussels, Belgium

**Keywords:** COVID-19, Excess Mortality, Linear Mixed Model, 5-year Weekly Average

## Abstract

The Corona Virus Disease (COVID-19) pandemic has increased mortality in countries worldwide. To evaluate the impact of the pandemic on mortality, excess mortality has been suggested rather than reported COVID-19 deaths. Excess mortality, however, requires estimation of mortality under non-pandemic conditions. Although many methods exist to forecast mortality, they are either complex to apply, require many sources of information, ignore serial correlation, and/or are influenced by historical excess mortality. We propose a linear mixed model that is easy to apply, requires only historical mortality data, allows for serial correlation, and down-weighs the influence of historical excess mortality. Appropriateness of the linear mixed model is evaluated with fit statistics and forecasting accuracy measures for Belgium and the Netherlands. Unlike the commonly used 5-year weekly average, the linear mixed model is forecasting the subject-specific mortality, and as a result improves the estimation of excess mortality for Belgium and the Netherlands.

## 1. Introduction

During the Corona Virus Disease (COVID-19) pandemic, most countries have reported the number of COVID-19 deaths as an essential part of their monitoring strategy (Giattino et al., 2021). However, reported COVID-19 deaths depend on the completeness and strategy of counting deaths. Variation in counting exists through testing strategy, availability of test material, in- or excluding nursing home deaths, or by variation in coding and registration. Hence, reported COVID-19 mortality is prone to mis-reporting. Therefore, excess mortality has been suggested to assess the overall impact on mortality of the SARS-CoV-2 virus (Aron et al., 2020; Morgan et al., 2020; Beaney et al., 2020). Excess mortality is obtained by subtracting the expected deaths based on the pre-pandemic period from the registered all-cause deaths in the pandemic period. Reported all-cause deaths are not only more reliable across countries, but excess death captures also both direct and indirect effects of a pandemic on mortality. Lower death counts due to mitigation measures or higher counts due to COVID-19 or an overloaded health system will both be reflected in excess mortality.

The critical part in determining excess mortality is a reliable estimate of *baseline* mortality, i.e., the mortality that is expected under non-pandemic conditions. A simple method to determine baseline mortality is the average of historical mortality data, most commonly the weekly average of the past 5 year, if available (Beaney et al., 2020; Michelozzi et al., 2020; Modig et al., 2020; Stang et al., 2020; Giattino et al., 2021). However, by predicting the population average, this methodology ignores the trend in mortality during the first weeks in 2020 before the pandemic, which was lower than average in, for example, Belgium (Molenberghs et al., 2020; Bustos Sierra et al., 2020). Additionally, the weekly average may be influenced by peaks of increased mortality due to heat waves or seasonal influenza epidemics in recent history. Two popular methods to minimize past influence of excess mortality in the forecasting of baseline mortality are the Farrington (Farrington et al., 1996) and Euro-MOMO models (Vestergaard et al., 2020; Fouillet et al., 2020). The Farrington model uses residuals to down-weigh the influence of outbreaks in the past, while the Euro-MOMO model takes only historical periods without excess mortality into account to forecast future baseline mortality. However, excluding Winter and Summer seasons due to influenza or heat waves, may not be sufficient to eliminate the influence of these events on mortality (Aron et al., 2020). The number of deaths in Spring may, for example, be below average after a severe seasonal influenza season.

Variations of the Euro-MOMO model exist that do not exclude historical data for mortality forecasting and add, similar to Serfling’s models (Serfling, 1963), a cyclic term to model seasonality (Cox et al., 2010; Nielsen et al., 2018, 2021). However, these extensions often require additional information, such as historical influenza data, temperature, ozone concentration, etc., which may not always be easy to access, especially not when several countries are considered in a common analysis. Time series models, such as Dynamic Harmonic Regression (Chen et al., 2020) and ARIMA models (Faust et al., 2021), exploit the serial correlation in the historical mortality data. The latter models however require stationarity of the time series and are excellent for short-term forecasting, but may be limited on the long-term horizon (Harvey, 1989; Harvey and Todd, 1983). Others have suggested simple linear models to determine excess mortality by COVID-19, including a yearly time trend in combination with fixed or smoothing spline weekly effects (The Economist, 2020; The New York Times, 2020). That said, the longitudinal mortality data likely violate the independent error assumption of these linear models.

We propose a linear mixed model based on uninterrupted historical mortality data to forecast the subject-specific baseline mortality for the year 2020 in order to estimate the COVID-19 excess mortality. While linear mixed models offer a versatile modeling family, which can incorporate many mean and (co)variability structures for longitudinal data, including serial correlation (Verbeke and Molenberghs, 2000; Verbeke et al., 1998; Chi and Reinsel, 1989), we show that they are particularly well designed for modeling baseline mortality patterns. The influence of historical excess mortality is downsized by two distinct strategies, by down-weighing the residuals, similar to Farrington et al. (1996), and by down-weighing the historical excess mortality data. Although marginal population average predictions and conditional subject-specific predictions cannot be compared directly, we will demonstrate the advantage of using the subject-specific predictions of the linear mixed model over the commonly used 5-year weekly population average in forecasting the mortality in 2020 for Belgium and the Netherlands.

## 2. Data

Open source daily all-cause mortality data of Belgium from the year 2009 onward are available from the national statistical institute, Statistics Belgium (STATBEL, 2020). These data were downloaded and temporally aggregated in weekly periods. The weeks are defined according to the International Standard ISO 8601 definition, i.e., Monday is the first day of the week and the first week of the year is the week which contains the first Thursday of January. The first week of the year 2009 was excluded from the data, since it is an incomplete week. Also, the weeks numbered 53, present in some years, are excluded.

In Belgium, daily COVID-19 mortality data are registered by Sciensano (EPISTAT: COVID-19, 2020). These open source data were extracted and aggregated in weeks using the same week definition as for the all-cause mortality. Registered COVID-19 related deaths in Belgium include confirmed and possible COVID-19 deaths (Bustos Sierra et al., 2020).

For the Netherlands, weekly historical mortality data are available from The Human Mortality Database (2021) for the year 1995 until 2020, while daily reported COVID-19 mortality is available from National Institute for Public Health and the Environment (2021). The same aggregation methods as used for the Belgian data are applied.

## 3. Model Proposal for Excess Mortality

In Section 3.1, a general linear mixed model is presented to forecast subject-specific mortality within a region, with a possible serial temporal correlation structure. In Section 3.2, two strategies for down-weighing past excess mortality, such as heat waves and influenza outbreaks, are added to the model. With these models the excess mortality is estimated in Section 4.

The data analyses were performed and figures produced using SAS 9.4 Software and R Studio 4.0.3.

### 3.1 Linear Mixed Model

We will model the weekly mortality *y*_*tj*_ with week *t* = 1, …, 52 by year *j* = 2009, …, 2020. For the year 2020, we will use only the first 10 weeks of the observed mortality and forecast the remaining weeks of the year. In Belgium, the first COVID-19 related death was reported in week 11 (Bustos Sierra et al., 2020), while 4 COVID-19 related deaths occurred in week 10 in the Netherlands. After a relatively mild influenza season during the Winter of 2019– 2020, Belgian and Dutch mortality counts during the first 10 weeks of 2020 were lower than average (Figure 1 and 2). This below-average mortality will influence expected mortality in the following weeks and thus should be taken into account when forecasting. Hence, we will focus on subject-specific predictions.

**Figure 1.**
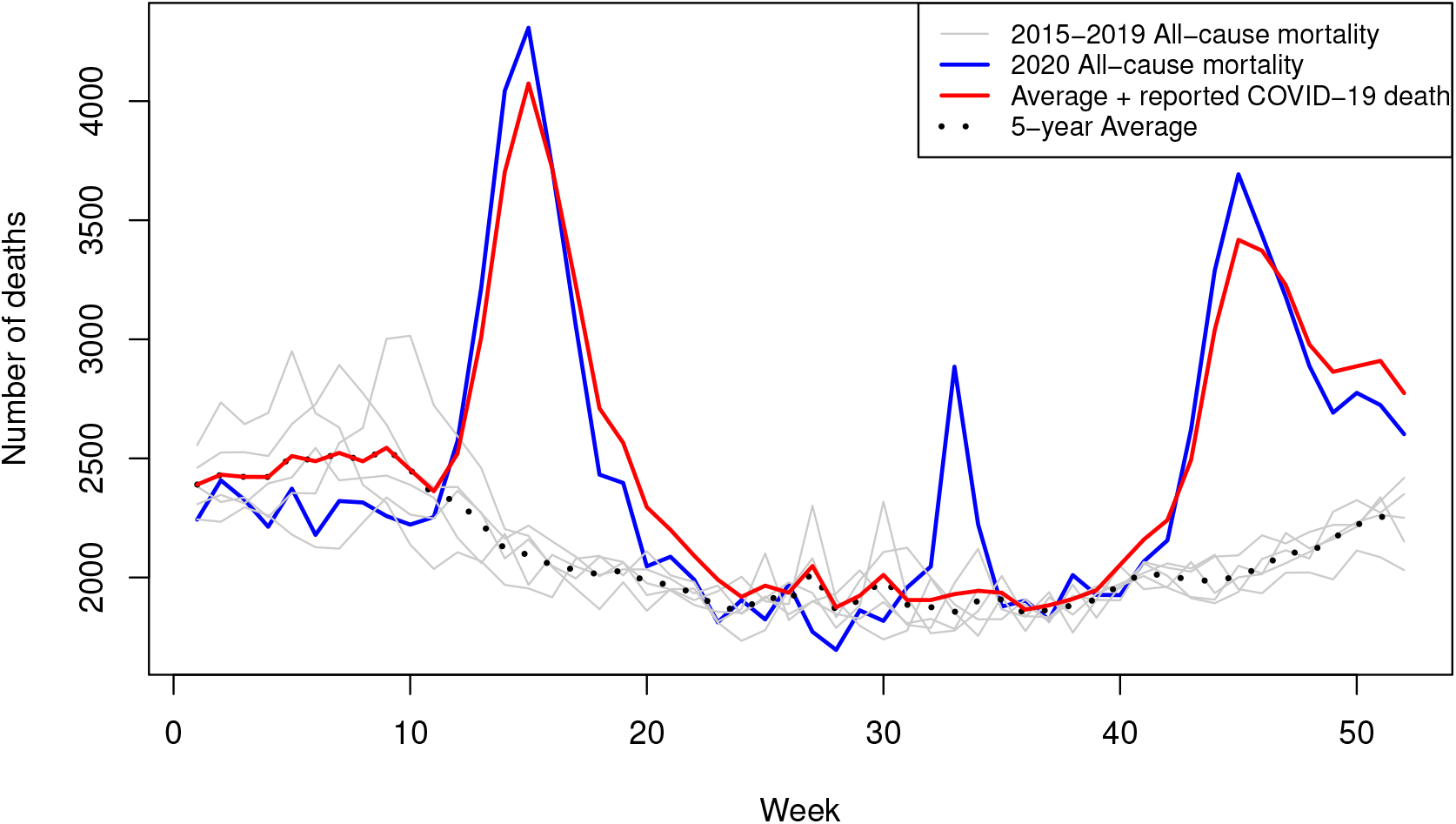
Weekly all-cause mortality in Belgium from year 2015 to 2020, with the 5-year average (years 2015–2019) and the sum of the 5-year average with the reported COVID-19 deaths.

**Figure 2.**
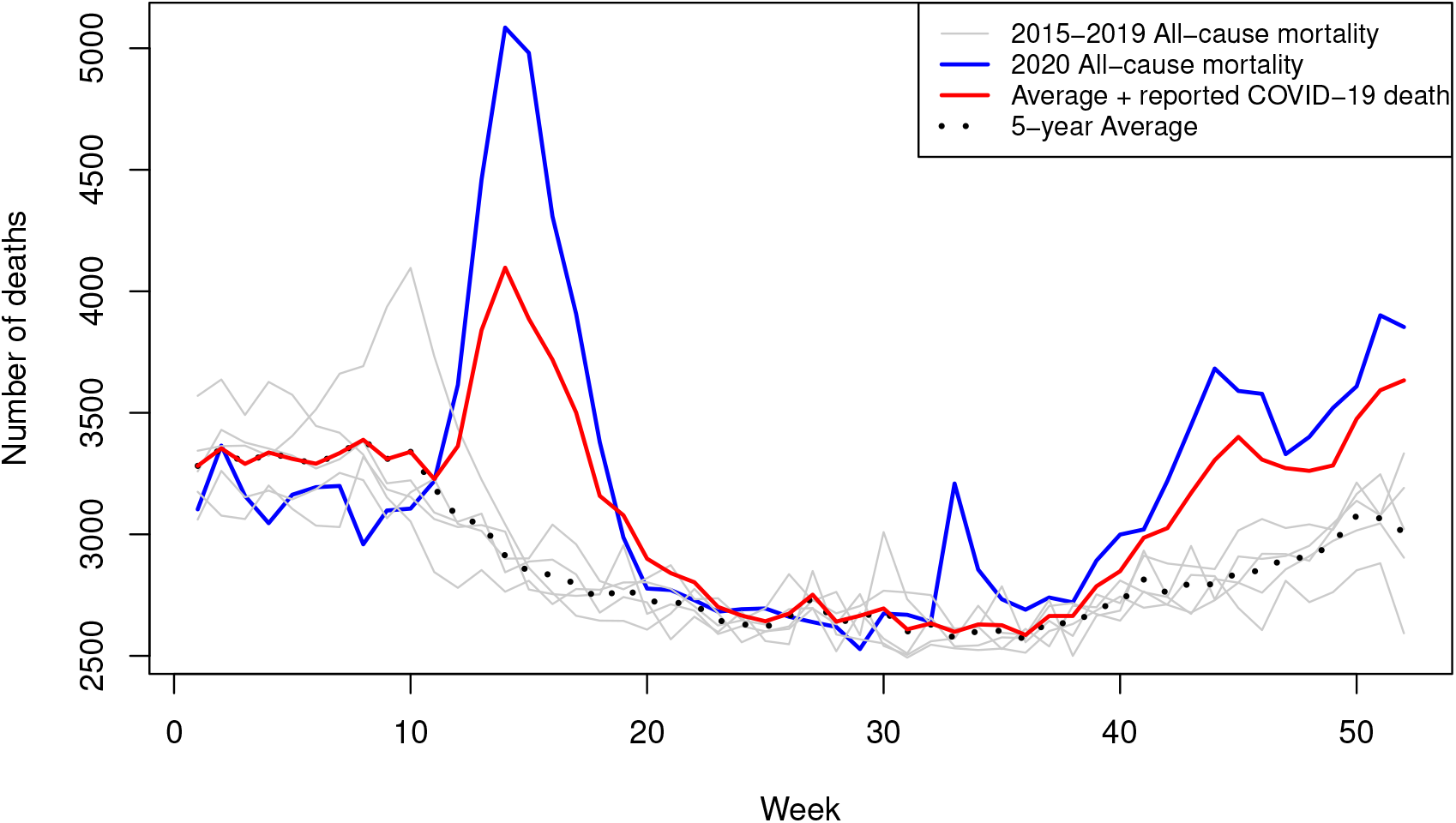
Weekly all-cause mortality in the Netherlands from year 2015 to 2020, with the 5-year average (year 2015–2019) and the sum of the 5-year average with the reported COVID-19 deaths.

The number of deaths is usually modelled with a Poisson distribution. However, since the mean of the weekly deaths is sufficiently high for the central limit theorem to be invoked, we will use a Gaussian model.

Mortality in Belgium and the Netherlands clearly shows a cyclic pattern (Figures 1 and 2). A correlogram indicates that a yearly cycle is strongly present with a less pronounced half-yearly cycle. Therefore, both yearly and half-yearly Fourier series are included into the model. As mortality may fluctuate year by year, for instance due to increasing population sizes or changing age distributions, a random intercept is added to the model. Additionally, variation in the cyclic pattern from year to year is allowed by including a random effect for the yearly sine wave, which results in the following model:

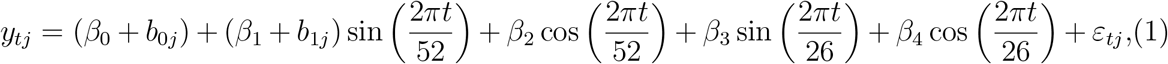

with *ε*_*tj*_ *∼ N* (0, *σ*^2^), **b**_**j**_ = (*b*_0*j*_, *b*_1*j*_) *∼ N* (0, *D*) and the *ε*_*tj*_ and **b**_**j**_ mutually independent.

Since it is known that random effects are often able to represent the serial correlation among the measurements (Chi and Reinsel, 1989; Verbeke and Molenberghs, 2000), and random-effects models including serial correlation may sometimes overparameterize the covariance structure, we will carefully evaluate via the likelihood ratio test if splitting the error in a serial correlation *ϵ*_(1)*t*_ and a measurement error *ϵ*_(2)*t*_ is required. In any case, it has been shown that, if serial correlation is present on top of the random effect correlation, the inclusion of a serial correlation structure is preferable over correctly specifying the model (Verbeke et al., 1998). The linear mixed model can be easily extended to include additional random effects, if needed. Model parameters are estimated via restricted maximum likelihood (REML), given that it outperforms ML estimators towards removing finite-sample bias (Molenberghs et al., 2020).

### 3.2 Reducing the influence of historical excess mortality

When estimating the baseline mortality, the influence on the parameter estimates of historical excess mortality, mainly due to heat waves and seasonal influenza epidemics, needs to be reduced. Two strategies are developed, which both require a 3-step analysis where model 3.1 is fitted twice.

The first method follows the weighted regression of Farrington et al. (1996) and downweighs historical excess mortality for standardized conditional residuals (Nobre and da Motta Singer, 2007), *r*_*tj*_ *>* 1. After fitting model (3.1), for the first time, a weight *w*_(1)*tj*_ ba sed on the standardized residuals *r*_*tj*_ is obtained:

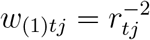

for *r*_*tj*_ *>* 1. Next, a weighted regression model 3.1 with weights *w*_(1)*tj*_ is fitted a second time.

The second method also uses the standardized residuals obtained after fitting model 3.1 a first time, but down-weighs the observations by multiplication with the weight:

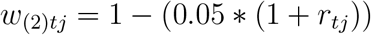

for *r*_*tj*_ *>* 1. Next, model (3.1) is fitted a second time, but now on the weighted observations. Observations in historical excess mortality weeks will have higher standardized residuals, which results in a larger reduction of the observation by the weight *w*_(2)*tj*_.

## 4. Estimating Excess Mortality

The weekly prediction and 95% confidence interval (CI) of the baseline mortality during the pandemic (week 11 to 52) are based on the year 2020 subject-specific conditional predictions of the linear mixed models. The weekly excess mortality results from subtracting the predicted mortality from the observed pandemic weekly mortality. Finally, the weekly estimated excess mortality, and similarly its lower and upper bounds, is summed over all pandemic weeks to results in an estimate and 95% CI of the excess mortality of the year 2020.

## 5. Application

The linear mixed model 3.1 is fitted to historical mortality data from week 2 of year 2009 to week 10 of year 2020 for Belgium and the Netherlands. The need for modelling additional correlation is evaluated by adding Gaussian serial correlation *ϵ*_(1)*t*_ to the measurement error or by adding an additional random effect *b*_3*j*_ on the half-yearly sine wave.

The appropriateness of the model is evaluated by several statistics. The likelihood ratio test compares the *™*2 log likelihood difference between model 3.1 and the expanded models with a mixture 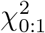 distribution for the serial correlation and mixture 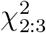 distribution for the half-yearly sine wave random effect. Additionally, the root mean square error percentage (RMSE%) evaluates the forecasting accuracy of the models. If the forecasting error *e*_*jt*_ is the difference between the forecasted death *f*_*jt*_ and observed death *y*_*jt*_, then:

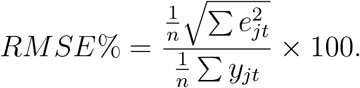

Since, the proposed models down-weigh the historical excess mortality, the forecasting accuracy measure can only sensibly be evaluated in years where there has been no or little excess mortality. Historical years with substantial excess mortality will by definition have a large deviation between the observed and forecast deaths for the weeks with excess mortality. For Belgium, influenza related mortality was very low in the year 2014. While 2016 had a heat wave with little excess mortality, both 2014 and 2016 had no marked episodes of higher mortality than expected. Both 2014 and 2016 will be evaluated by excluding deaths from week 11 onward from these years during estimation and forecast the baseline mortality using the first 10 weeks.

For Belgium, adding either Gaussian correlation or an additional random effect for both the weighted regression as the weighted observations strategy significantly improves the model (Table 1), although the estimated excess mortality from week 11 to 52 for the year 2020 is not very different between all the models. Other serial correlation structures either do not converge or fit the data significantly worse. For the weighted regression models, the model including the Gaussian serial correlation fits the data best, while for the weighted observations, the model with the two random sine wave effects fits the data slightly better. Comparing the log-likelihood between the weighted regression and weighted observation models is inappropriate given that the observations between the two models are distinct.

**Table 1.**
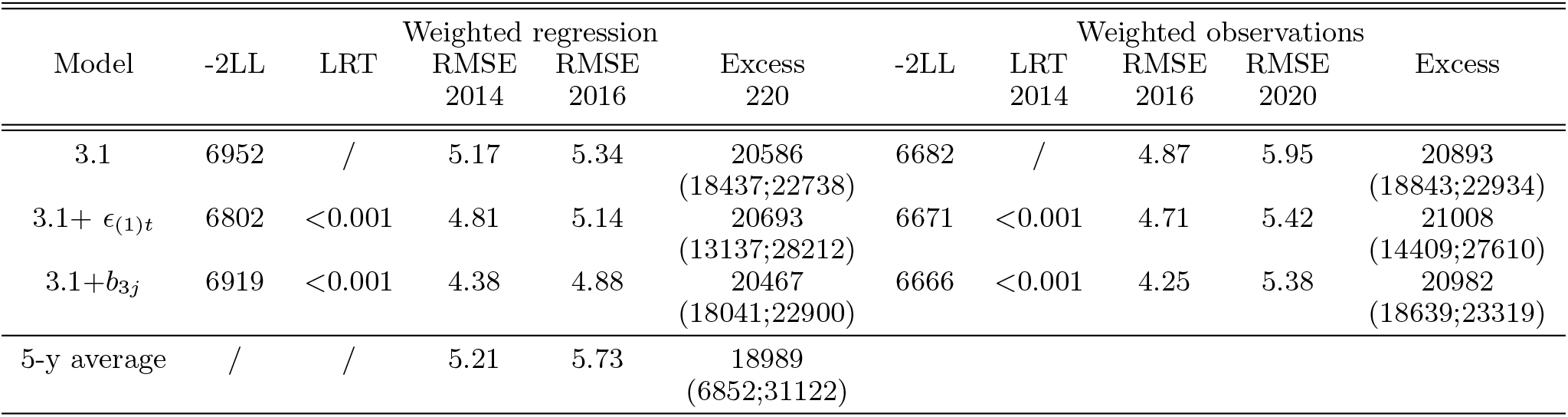
Model fit, forecasting accuracy and excess mortality estimation (95% CI) comparing Linear mixed models and 5-year weekly average method for Belgium. LL=log likelihood, LRT=Likelihood ratio test and RMSE=root mean square error.

The forecasting accuracy of the subject-specific linear mixed model in Belgium for both years 2014 and 2016 is better than the 5-year weekly population average (Table 1). Note that for the 5-year weekly average, the years 2009–2013 were used to forecast mortality for 2014 and the years 2011–2015 to forecast 2016. Although the differences are small, the model with two random sine wave effects has a slightly better forecasting accuracy. Comparing the weighted regression models with the weighted observation models, there is little difference in forecasting accuracy. For the year 2014 the weighted observation models have slightly better forecasting accuracy, while it is the reverse for 2016. Using the Mean Absolute Error (MAE) or Mean Absolute Percentage Error (MAPE) as forecasting accuracy measure, the conclusions remain the same.

A clear advantage of the linear mixed models over the 5-year weekly average is the accuracy in estimation of the baseline and excess mortality. The 95% confidence interval of the mortality forecasting is much wider for the 5-year weekly average (Table 1). Using the years 2009 to 2019, rather than only the last 5 years, decreases the variability for the 5-year average (excess mortality estimate: 19,957 with 95% CI 10,531;29,381), but it is still wider then the linear mixed model variance. When using only the last 5 years to fit the linear mixed models, convergence issues arise because of insufficient data to estimate the correlation structure.

From week 11 to 52 in 2020, 19,288 COVID-19 deaths were reported in Belgium (EPISTAT: COVID-19, 2020). Adding the excess mortality of 1460 deaths from the heat wave during the Summer of 2020 (Bustos Sierra et al., 2020), it seems that the linear mixed model forecasts excess mortality well (Figure 1 and Table 1).

In the Netherlands, the first COVID-19-related death was reported on March 6, 2020 (National Institute for Public Health and the Environment, 2021). Since only 4 deaths were reported in week 10, we chose also for the Netherlands to initiate forecasting mortality from week 11 onward. In the Netherlands the year 2020 started with a lower-than-average mortality during the first 10 weeks (Figure 2). The likelihood ratio tests show that adding a correlation structure to model 3.1 significantly improves the model fit for the historical mortality data in the Netherlands (Table 2). Although the variability between the estimation of the excess mortality of the different models is larger than in the Belgian analysis, the precision of each estimation is much better than the 5-year weekly average. Similar to Belgium, only data from 2009–2019 have been used to fit the linear mixed models. Using data from 1995 onward for the Netherlands does not reduce the variance of the excess estimation by the linear mixed models much.

**Table 2.** Model fit and excess mortality estimation (95% CI) comparing Linear mixed models and 5-year weekly average method for the Netherlands. LL=log likelihood and LRT=Likelihood ratio test.

| Model | Weighted regression |  |  | Weighted observations |  |  |
| --- | --- | --- | --- | --- | --- | --- |
|  | -2LL | LRT | Excess 2020 | -2LL | LRT | Excess 2020 |
| 3.1 | 7109 | / | 20025<br>(15634;24414) | 7020 | / | 21125<br>(16663;25589) |
| 3.1+$\epsilon_{(1)t}$ | 6935 | <0.001 | 20698<br>(11534;29858) | 7001 | <0.001 | 20727<br>(11337;30113) |
| 3.1+$b_{3j}$ | 7044 | <0.001 | 20585<br>(15023;26145) | 7004 | <0.001 | 22796<br>(17308;28277) |
| 5-y average | / | / | 19024<br>(5324;32726) |  |  |  |

Contrary to Belgium, in the Netherlands the reported COVID-19 deaths (11,527 across weeks 11–52 in 2020) is clearly less than the estimated excess mortality by both the linear mixed models and 5-year weekly average (Figure 2 and Table 2). Taking account of the heat wave in the Netherlands in week 33, which resulted in an estimated 400 excess deaths (National Institute for Public Health and the Environment, 2021), the linear mixed models estimate that between 51–56% of the COVID-19 mortality has been reported in 2020.

## 6. Discussion

In 2020 the world was confronted with the most lethal pandemic in one hundred years, the severity of which is underscored by the all-cause mortality from Belgium and the Netherlands. They show how hard we were hit, but also how important the mitigation measures against the spread of the virus are. To understand the complete picture of the pandemic, for individual countries and for the comparison between countries, it is useful to estimate the excess mortality, since both direct and indirect effects of the pandemic on mortality are captured by excess mortality.

Determining excess mortality requires the estimation of mortality under non-pandemic conditions. The often used method of averaging the 5-year historical weekly mortality, however, ignores the trend in mortality of the first weeks of 2020 by estimating the population average and may be influenced by recent excess mortality in the past. The advantage of the weekly average method is that it is easy to apply and only information about mortality is required. We propose linear mixed models to forecast subject-specific baseline mortality, which address the shortcomings of the 5-year weekly averaging method, while maintaining simplicity in application and limited requirements of data. Indeed, as compared to time series models, the linear mixed models require less expertise to fit (Harvey, 1989), but also result in a smaller variance for the forecast values.

For Belgium and the Netherlands, we have shown that the linear mixed models not only fit the mortality data better, but also that the prediction for the years 2014 en 2016 are superior to the 5-year weekly average. For Belgium, the excess mortality in 2020 is estimated by the linear mixed models to lie between 20,467 and 21,008. Taking account of 1460 excess deaths during the Summer heat wave in 2020 (Bustos Sierra et al., 2020), between 19,000 and 19,550 deaths can then be attributed to direct and indirect effects of the COVID-19 pandemic. Since 19,288 COVID-19 related deaths have been reported in Belgium (EPISTAT: COVID-19, 2020), the country has reported COVID-19 mortality fairly well. For the Netherlands, the excess mortality in 2020 is estimated by the linear mixed models to lie between 20,585 and 22,796, although only 11,527 COVID-19 deaths have been reported (Ritchie et al., 2021). Likely, the Netherlands have under-reported COVID-19 deaths by an estimated 51–56%. Recently, the Dutch Central Bureau of Statistics (CBS) have attributed a little more than 20,000 deaths in 2020 to COVID-19 (Central Bureau for Statistics, 2021), which is in line with the estimation by the linear mixed models.

Both 2020 waves of SARS-CoV-2 infections in both Belgium and the Netherlands have led to thousands of COVID-related deaths. While the first wave was shorter and more intense, the second wave, which started roughly at week 30 and continued after the study period investigated here, was longer, resulting in more or less equal numbers of COVID-19 deaths in both periods. As the timing of the deaths are parallel to the excess mortality, it shows that likely not the non-pharmaceutical interventions, but the virus itself is responsible for the majority of the excess mortality.

It is of course artificial to evaluate the effect of the pandemic on mortality by calendar year. The full impact of COVID-19 on mortality will be evident when sufficient individuals in the population will be vaccinated and the number of COVID-19 deaths greatly reduced. This would also allow for international comparison between countries with a different timing of the pandemic or with different mitigation measures. In the meantime, using the linear mixed models allow for intermediate evaluation.

Finally, excess mortality over an entire country does not tell the complete tale of the effect of the COVID-19 pandemic on mortality. Age groups, gender and regional difference within a country are smoothed out in an overall excess-mortality value. To account for these differences, the linear mixed models can be extended to include these variables or can be applied to the different subgroups. They can also be applied to historical data that are interrupted, for example if only seasonal historical mortality data are available.

## Data Availability

The data is openly available

## Acknowledgments

We are grateful for the ability to use the historical mortality data (Statbel, Belgium and the Human Mortality Database, The Netherlands) and the data on COVID-19 confirmed deaths (Sciensano, Belgium, RIVM, The Netherlands). The data providers hold no responsibility for the analyses reported in this manuscript. *Conflict of Interest*: None declared.

